# DoBSeqWF: A framework for sensitive detection of individual genetic variation in pooled sequencing data

**DOI:** 10.1101/2025.04.23.25326275

**Authors:** Mads Cort Nielsen, Christian Munch Hagen, Ulrik Kristoffer Stoltze, Thomas van Overeem Hansen, Mette Nyegaard, Henrik Hjalgrim, Marie Bækvad-Hansen, Anna Byrjalsen, Kjeld Schmiegelow, Karin Wadt, Jonas Bybjerg-Grauholm, Simon Rasmussen

## Abstract

**Motivation:** Population screening for rare genetic diseases is limited by the high cost of next-generation sequencing. Double-batched sequencing (DoBSeq) is a cost-effective method for assigning rare variants to individuals using two-dimensional unique double- pooled sequencing. However, this method produces complex, high-depth sequencing data that requires a specialized workflow for efficient and reproducible analysis.

**Results:** We developed DoBSeqWF (DoBSeq Workflow), a Nextflow-based pipeline for processing the pooled sequencing data from alignment through variant calling, filtering, and ultimately individual assignment of rare variants. Using separate training and validation datasets with whole genome sequencing as the gold standard, we benchmarked multiple variant callers, and we developed and implemented machine learning filters that improve rare variant calling performance while maintaining high sensitivity. The pipeline enables reproducible analysis and can be easily updated as bioinformatic tools and variant interpretations evolve.

**Availability and Implementation:** DoBSeqWF is freely available at https://github.com/RasmussenLab/DoBSeqWF.

## Introduction

Rare diseases (RDs) are individually rare but accumulate to affect a large proportion of the population. Depending on the definition, as many as 10,000 RDs have been identified (Haendel *et al*. 2020), and it is estimated that at least 6% of the European population suffers from some RD (Ferreira 2019). The consequences of living with an RD include delayed or mis-diagnosis, incorrect treatment, prolonged hospitalisation and increased mortality (Bick *et al*. 2019). In addition to affecting the individual, it indirectly impacts society by increasing healthcare costs and causing loss of work ability (Chung *et al*. 2022). The low prevalence and lack of identifiable biomarkers for many RDs render them unsuitable for diagnostic testing using traditional biochemical assays at scale (Stark and Scott 2023).

However, at least 70% of RDs have a defined genetic etiology, and many are caused by pathogenic variants in single genes (Amberger *et al*. 2015; Boycott *et al*. 2017). This makes testing using genetic sequencing a promising approach, which can include hundreds of RDs in a single test (Wright, FitzPatrick and Firth 2018). Arguably, the high cost of sequencing has been a main factor preventing the widespread use of genetic testing for screening. In a recent pilot study, we addressed this with a novel approach termed DoBSeq (double-batched sequencing), reducing the cost of genetic testing of rare variants to a fraction compared to individual sequencing (Stoltze *et al*. 2023). The method successfully implements pooled double sequencing, where any two pools overlap by only one individual (Zuzarte *et al*. 2014; Furstenau *et al*. 2020). In this method, individual samples of DNA are pooled, sequenced, and analyzed twice with overlap, making it possible to pinpoint or assign unique rare variants to their unique host. In this context, pinpointable rare variants (to ensure they are carried by only one individual in a pool) are defined as variants that are private or unique to the individual within the matrix.

DoBSeq mainly reduces costs by reducing the library kit usage and the number of samples handled after the initial pooling step. Depending on the number of samples in each pool, sequencing is required to a high depth (∼2,000X for 100 samples with 10 in each pool) to cover each allele for confident variant calling. The resulting pooled sequence data samples have a high allele count and depth, and in contrast to individual sequencing data, no established best practices exist for such variant analysis.

We hypothesize that the increased data complexity leads to a possible decrease in variant calling accuracy and that careful considerations should be made to choose the most suitable variant caller. Furthermore, many variant callers are designed to be lenient, requiring manual filtration, hard quality thresholds, or automated filters (Koboldt 2020). False positive variant calls are typically based on sequencing artifacts arising from the library preparation process or the sequencing itself and depend on both the sample preparation method and sequencing platform (Li 2014; Das, Biswas and Basu 2023). This inherent complexity of the errors, combined with the repeated data generation with screening, makes it a suitable case for training a filtering model that improves over time as more data is generated.

Here, we present an automated workflow for analyzing DoBSeq data and a variant filtering module based on machine learning. Our pipeline implements an approach for processing overlapped pooled sequencing data, from raw reads to variant assignment.

We benchmark multiple variant callers using a cohort of childhood cancer patients with available whole genome sequencing data as gold standard and develop a machine learning-based filtering module that outperforms traditional filtering methods. Our results demonstrate that the DoBSeq approach can reliably detect rare variants while reducing sequencing costs by 80-90 percent compared to individual testing. The automated workflow enables reproducible analysis and easy adaptation as variant databases and interpretation evolve, making it a promising tool for cost-effective population-level genetic screening.

## Methods

### Cohort

To optimize and validate the DoBSeq pipeline, we included samples from the Danish childhood cancer genomics study STAGING. The same subset and sequencing data previously described in Stoltze et al. (2023) were used. Individuals from the cohort were divided into two separate 10×10 DoBSeq matrices, each containing 100 individuals, which served as a training and held-out test set for the study.

### Individual whole-genome sequencing data and variant calls

Individual whole-genome NGS data was generated from leukocytic DNA using the HiSeqX platform (Illumina, San Diego, CA, USA) with paired-end sequencing of 150bp reads with a target average coverage of 30X. Raw reads were aligned to the GRCh38 primary human reference genome sequence using BWA-MEM2 (Li and Durbin 2010) (v. 2.2.1). The aligned reads were then preprocessed to remove duplicate read pairs and for base quality score recalibration using GATK (McKenna *et al*. 2010) (v. 4.3.0) according to GATK Best Practices (DePristo *et al*. 2011; Van der Auwera *et al*. 2013). Finally, the processed reads were subset to the genomic intervals of the 113 genes covered in Illumina’s TruSight Hereditary Cancer Panel (http://www.illumina.com/TruSightHereditaryCancer) using SAMtools v. 1.18 (Danecek *et al*. 2021). GATK joint genotyping was performed by first calling variants for the individual samples using HaplotypeCaller in GVCF mode. Individual VCF files were combined into multi-sample VCFs separately for the training and test sets using GenomicsDB import, and the final calls were generated using GATK GenotypeGVCF. Additionally, variants were called based on the preprocessed reads using DeepVariant (Poplin *et al*. 2018) (v. 1.5.0) in WGS mode using default settings. Using bcftools (Danecek *et al*. 2021) (v. 1.16), the variants in the resulting VCF files were normalised by ensuring concordance of reference alleles with the reference genome, indels were left-aligned, and multi-allelic variant sites were split. No additional filtering was performed, and the intersection and symmetric difference of variant calls between GATK and DeepVariant were then defined as high and low confidence, respectively.

### Overlapped pooled sequencing data

The laboratory protocol of DoBSeq is described in detail in Stoltze et al. (2023). In brief, for each training and test set, DNA was extracted from two 3.2-mm discs stamped from at-birth Guthrie cards for each individual. DNA concentrations were measured and normalized to ensure equimolar contributions from each sample. DoBSeq arranged the normalized DNA from the 100 individuals in a two-dimensional 10×10 matrix. The DNA was then pooled twice, once along each dimension, resulting in the DNA being present in exactly two pools: one along the rows and one along the columns (see **Figure 1**). The 20 pools were sequenced using the TruSight Hereditary Cancer Panel, which covers 113 genes commonly associated with cancer predisposition, on the NextSeq 500 platform with a 150bp paired-end sequencing kit and a target average coverage of 2,000X per pool.

**Figure 1.**
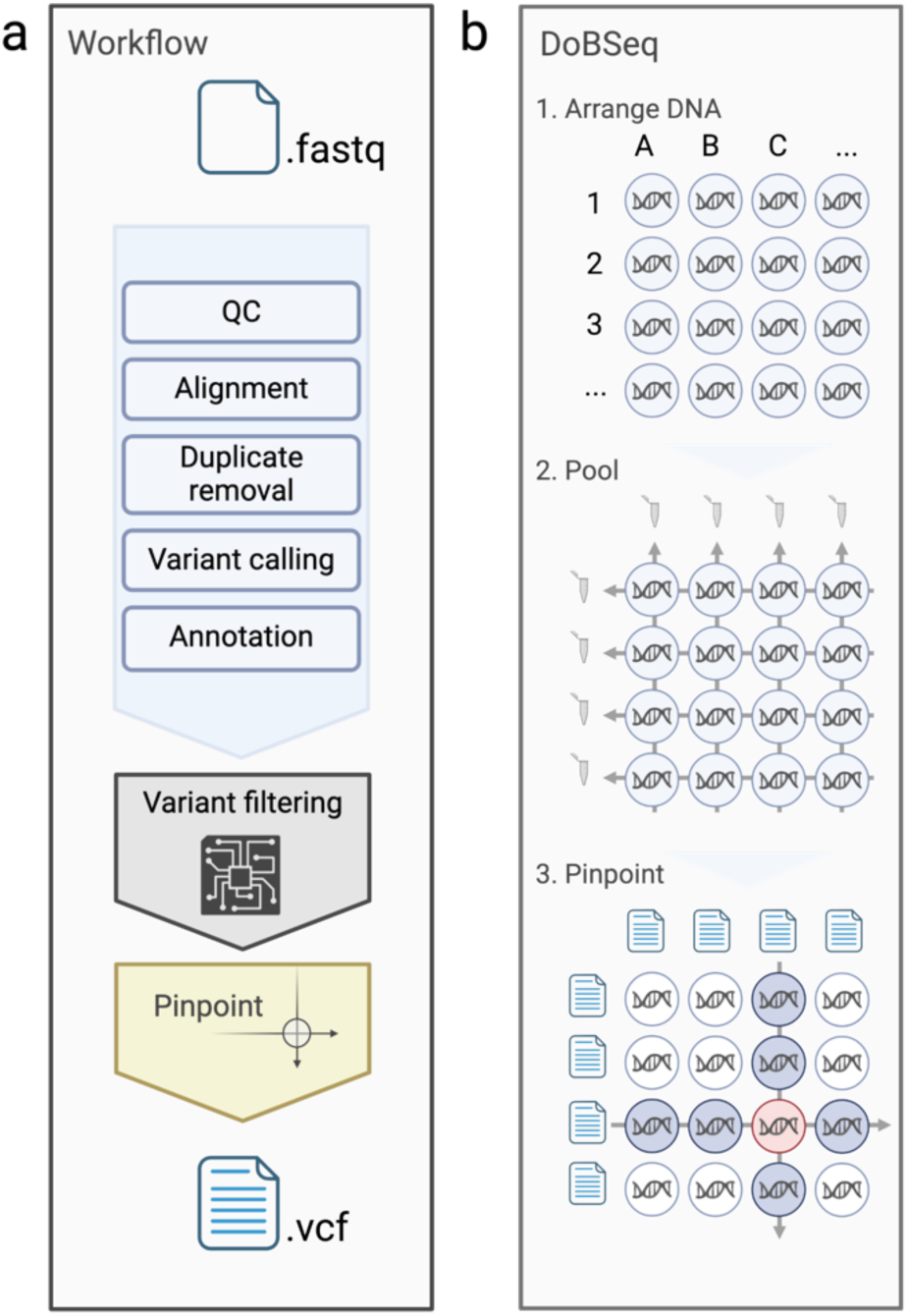
Pipeline for assigning rare variants to individuals in pooled sequencing data. **(a)** The DoBSeq workflow processes pooled sequencing data through quality control, alignment, duplicate removal, variant calling, (optional) machine learning-based filtering, and variant pinpointing, outputting VCF files with assigned variants. **(b)** The DoBSeq method: 1) DNA samples are arranged in a matrix; 2) DNA is pooled along rows and columns and sequencing follows; 3) Rare variants are pinpointed (assigned) to individuals by identifying variants present in exactly one row pool and one column pool, corresponding to a unique individual in the matrix.

### Analysis of pooled sequencing data

Using the DoBSeq pipeline developed in this study (v. 0.2.0), the input dataset of FASTQ files containing pooled sequencing data underwent an initial quality control using FastQC (Andrews 2010) (v. 0.11.8). Similar to the individual “gold standard”, the data was aligned to the GRCh38 primary human reference genome using BWA-MEM2 (Vasimuddin *et al*. 2019) (v. 2.2.1) and then preprocessed to remove duplicate read pairs using GATK MarkDuplicates (McKenna *et al*. 2010) (4.6.0.0). Variant calls were generated from the processed reads using GATK Haplotypecaller (McKenna *et al*. 2010) (4.6.0.0) in GVCF mode with allele-specific annotation output, following either individual genotyping using GATK GenotypeGVCF (McKenna *et al*. 2010) (4.6.0.0) or with an intermediary merging of GVCF files using CombineGVCF (McKenna *et al*. 2010) (4.6.0.0) for joint genotyping. Furthermore, three different specialized variant calling tools were used: CRISP (Bansal 2010) v. 0.2, LoFreq (Wilm *et al*. 2012) v. 2.1.3.1, and Octopus (Cooke, Wedge and Lunter 2021) v. 0.7.4. All tools were run using the correct ploidy (=20) or pool size settings and the proper platform-specific error model if applicable. In addition to the default runs, variant calls were also generated for GATK individual genotyping without QUAL filtering by setting “-stand-call-conf” to zero, and similarly for LoFreq by removing default filters using the arguments “--no-default-filter” and increasing the the P-value to 1 with “--sig 1”. Similar to the gold standard calls, variants were checked, indels were left-aligned, and multi-allelic variant sites were split using bcftools norm.

### Variant pinpointing

A custom Python script was developed based on the method described in detail in Stoltze et al. (2023) to pinpoint variants to individuals in the matrix (see **Figure 1**). The algorithm iterates through the matrix one element (individual) at a time. At each step, it performs set operations to identify variants found uniquely in the current row compared to all other rows and variants found uniquely in the column compared to all other columns. The variants common between unique column and row variants were then defined as pinpointables.

### Dataset and feature selection

The dataset used for developing the variant filtering model consisted of all variant calls generated by the GATK individual genotyping of pools in the training matrix. Each variant call was treated as a sample and labeled as either 0 (discordant) or 1 (concordant) based on its agreement with the low-confidence calls in the gold-standard individual call set for the specific pool. Multiple feature subsets were evaluated based on all available GATK technical annotations. One subset was created by identifying and removing highly correlated features, by calculating, clustering and plotting correlation coefficients using the R package corrplot (Wei and Simko 2024). Three additional subsets of 5, 10, and 15 features were generated using the Minimum Redundancy Maximum Relevance (MRMR) algorithm (Radovic *et al*. 2017). A final subset included only normally distributed features (see **Supplementary Table 1**).

### Machine learning models for variant filtering

Four machine-learning approaches were evaluated for variant filtering: Gaussian Mixture Models (GMM), Random Forest (RF), Logistic Regression (LR), and Gradient Boosting (XGB). The first three were implemented using scikit-learn (Pedregosa *et al*. 2011) v1.6.1, while XGB was implemented with XGBoost (Chen and Guestrin 2016) v2.1.3. Models were trained and evaluated separately on single nucleotide variant (SNV) and insertion/deletion (indel) data using repeated nested cross-validation on the training matrix (5 repetitions, 10 outer folds and 5 inner folds). A range of hyperparameters, as well as the different feature sets, were explored during the training and validation (see **Supplementary Table 2**). Based on the predicted probabilities in the training set, two prediction thresholds were established for each model: one optimizing the F1 score and another targeting 99% sensitivity. Final models were trained using the optimized hyperparameters on the full training matrix and evaluated on a separate held-out test matrix (see **Supplementary** Figure 1).

### Benchmarking filtration performance

Variant filtering using GATKs variant quality score recalibration (VQSR) was performed on the jointly genotyped training dataset using a range of tranche values (90-99.9) for SNVs and indels. The best-performing tranche values, based on the F1-score, were then used for the test set. Hard threshold levels were based on the parameters defined in the GATK best practices (DePristo *et al*. 2011; Van der Auwera *et al*. 2013). A variant would be characterized as a true positive (TP) if it was called in any of the gold standard individual WGS call sets for the specific pool and as a false positive (FP) if it was not. If a variant was found in any WGS call sets but not in the pool, it would be characterized as a false negative (FN). Performance metrics would then be estimated as follows:

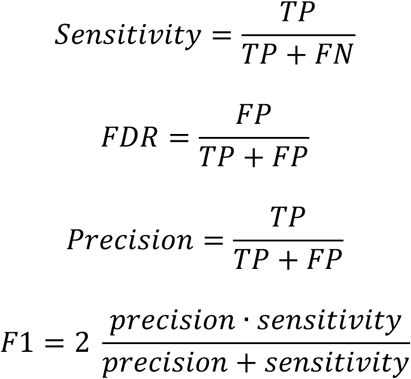

For the predictions in the held-out test set, 95% confidence intervals for each metric were estimated by bootstrapping (n=1000) and calculating the 2.5% and 97.5% percentiles from the resulting distributions.

## Results

### Pipeline for variant calling in overlapped pooled sequencing data

The DoBSeq workflow was developed to enable efficient, scalable, reproducible analysis of overlapped pooled sequencing data. It was implemented in Nextflow domain-specific language 2 (Di Tommaso *et al*. 2017). For the initial sequence analysis steps, based upon GATK best practices for germline variant calling while introducing a reference-free filtering module and a variant assignment algorithm. The pipeline consisted of four main steps: mapping, calling, filtering, and pinpointing (**Figure 1**). The input for the pipeline was a set of NGS FASTQ files and a sample table with information on matrix organization. The mapping step performs raw sequence quality control, alignment, removal of duplicate reads, and a range of optional alignment quality checks. Variant calling was done by GATK Haplotypecaller with allele-specific annotation output.

The optional filtering module developed in this study, then processes and filters likely false positives in the VCF files using an SNV-specific logistic regression and an indel- specific random forest model. Variants are then assigned to each individual using the DoBSeq algorithm, which identifies unique variants in the overlapping pools, resulting in VCF files with pinpointable variants as output. The modular design allows independent execution of each step, reducing computational overhead when optimizing parameters or updating reference files. The pipeline includes optional modules for variant annotation and report generation. The fully documented pipeline is available at https://github.com/RasmussenLab/DoBSeqWF.

### Generation of a gold standard dataset from individual WGS data

To optimize and validate the DoBSeq pipeline, we used two separate datasets, a training set and a held-out set, each composed of a 10×10 DoBSeq matrix of 100 individuals with WGS data available. To estimate the true genetic variation within the experiment target regions of a 113-gene panel, we used individual WGS data processed according to GATK best practices, and variants called using the two state-of-the-art germline variant callers HaplotypeCaller and DeepVariant. The outputs of the two variant callers were similar but not identical. For the training set, the callers had a concordance of 14,887 variants, which we labeled high confidence and 1273 variants called by only one of them, labeled low confidence. The absolute number of low confidence variants was similar for SNVs and indels (588/685; SNVs/indels), but because of the much lower total number of indels, the low confidence accounted for 32% (685/2126, low confidence/total). The majority of low confidence indels (71%, 489/685) were called by HaplotypeCaller, indicating that DeepVariant was, in agreement with Lin et al. (2022), more conservative. According to the DoBSeq matrix design, the variants of individuals were combined into pools by columns and rows and annotated by their theoretical ability to be pinpointed by the DoBSeq method. This aggregation resulted in a final set of 20 theoretical pools, with an average of 742.5 total and 60 pinpointable high-confidence variants per pool. This created a validation dataset with estimates of the true variant content and the correct individual assignments. The same analysis was done for the held-out matrix.

### Variant calling in DoBSeq overlapped pooled sequencing data

Unlike germline variant calling in individual NGS data, no well-established guidelines exist for calling variants in overlapped pooled sequencing data. To optimize the DoBSeq pipeline, we evaluated multiple variant callers on the training matrix against the high confidence validation dataset: GATK (widely used in clinical settings), LoFreq (optimized for low-frequency variants), Octopus (optimized for polyploid data), and CRISP (designed for pooled sequencing). Several technical limitations emerged during testing. Octopus failed to analyze two of the 20 pools due to RAM limitations (>1600GB required), while GATK joint genotyping required manual splitting of genomic regions due to Java array length limitations. However, all callers achieved high sensitivity (0.88-0.96, see **Table 1**), with GATK joint genotyping showing the lowest sensitivity (0.88) and LoFreq the highest (0.96). GATK individual genotyping had the highest F1 score, indicating a balance between sensitivity and FDR. An attempt to increase the sensitivity of the two best candidates, LoFreq and GATK individual genotyping, by reducing call confidence thresholds had varying effects: GATK maintained a high F1 score (0.85) with increased sensitivity (0.96), while LoFreq’s false discovery rate increased from 0.29 to 0.75. Notably, CRISP, despite being specifically designed for pooled sequencing, showed no performance advantage over other callers (F1: 0.84, sensitivity: 0.93). Our results indicate that specialized variant callers did not outperform GATK for pooled sequencing data, leading us to select GATK individual genotyping as our default caller.

**Table 1.** Comparison of variant callers for two-dimensional overlapping pooled sequencing data. Variants were called in pools of 10 individuals in a 10×10 matrix using four variant callers. Variant calling was performed using lowered (lenient) confidence thresholds for GATK and LoFreq in addition to default parameters. All calls, common and rare, in all pools, were compared to the intersection of GATK and DeepVariant calls in individual WGS data. *Octopus failed to analyze two of the 20 pools.

| Calling method | Total | TP | FP | FN | Sensitivity | F1 | FDR |
| --- | --- | --- | --- | --- | --- | --- | --- |
| GATK | 14887 | 14206 | 3534 | 681 | 0.954 | 0.871 | 0.199 |
| GATK lenient | 14887 | 14335 | 4385 | 552 | 0.963 | 0.853 | 0.234 |
| GATK joint | 14887 | 13126 | 4077 | 1761 | 0.882 | 0.818 | 0.237 |
| CRISP | 14887 | 13904 | 4209 | 983 | 0.934 | 0.843 | 0.232 |
| LoFreq | 14887 | 14332 | 5866 | 555 | 0.963 | 0.817 | 0.29 |
| LoFreq lenient | 14887 | 14345 | 41619 | 542 | 0.964 | 0.405 | 0.744 |
| Octopus* | 13399* | 12799 | 2551 | 600 | 0.955 | 0.890 | 0.166 |

**Table 2.** Performance of variant calling and filtering approaches on pinpointable variants. Performance before and after filtering variant calls from GATK individual genotyping (ML-S/F1, hard filtering thresholds) or joint genotyping (VQSR) workflows. *The joint genotyping workflow failed to analyse all pools.

| Calling method | Filtering approach | TP | FP | FN | Sensitivity | FDR | F1 |
| --- | --- | --- | --- | --- | --- | --- | --- |
| GATK | No filtering | 619 | 74 | 61 | 0.910 | 0.123 | 0.893 |
| GATK joint* | No filtering | 617 | 57 | 63 | 0.907 | 0.085 | 0.911 |
| GATK | Hard threshold | 3 | 3 | 677 | 0.004 | 0.571 | 0.009 |
| GATK | ML - F1 optimised | 590 | 27 | 90 | 0.868 | 0.045 | 0.909 |
| GATK | ML - sensitive | 612 | 52 | 68 | 0.900 | 0.081 | 0.909 |
| GATK joint* | VQSR | 597 | 30 | 83 | 0.904 | 0.070 | 0.917 |

### A machine learning method for removing false positive variant calls

Even with the best-performing variant calling method, false positive calls accounted for almost 20% in the pooled sequencing data. This high error rate confirmed the need for additional filtering to avoid false variant assignments and missed true variants due to false calls in overlapping pools. To address this challenge, we evaluated four different machine-learning approaches: Gaussian mixture models (GMM), random forest (RF), gradient boosting (GB), and logistic regression (LM). Models were trained separately for SNVs and indels using technical annotations from GATK as features and validated using repeated nested 5-fold cross-validation (see **Supplementary** Figure 2).

Performance was assessed using F1-optimized and high-sensitivity (99%) thresholds on the held-out test matrix. All models performed well in filtering SNVs, achieving high sensitivity (0.98-0.99) and low false discovery rates (0.04-0.08) with F1-optimized thresholds (see **Figure 2**). The logistic regression model slightly outperformed other approaches for SNVs with the highest F1-score of 0.974 (95% CI: 0.973-0.976) and lowest false discovery rate of 0.043 (95% CI: 0.039-0.046), while GMM showed relatively lower performance with an F1-score of 0.951 (95% CI: 0.948-0.954) and a higher FDR of 0.077 (95% CI: 0.073-0.082). All models achieved high sensitivities exceeding 0.98. However, indel filtering proved more challenging across all models, with sensitivities ranging between 0.65 (95% CI: 0.62-0.68) for logistic regression and 0.74 (95% CI: 0.71-0.77) for random forest, and widely varying false discovery rates from 0.28 (95% CI: 0.25-0.31) for XGBoost to 0.71 (95% CI: 0.70-0.73) for GMM. Based on these results, we implemented a combined approach using logistic regression for SNVs and random forest for indels in the final workflow module.

**Figure 2:**
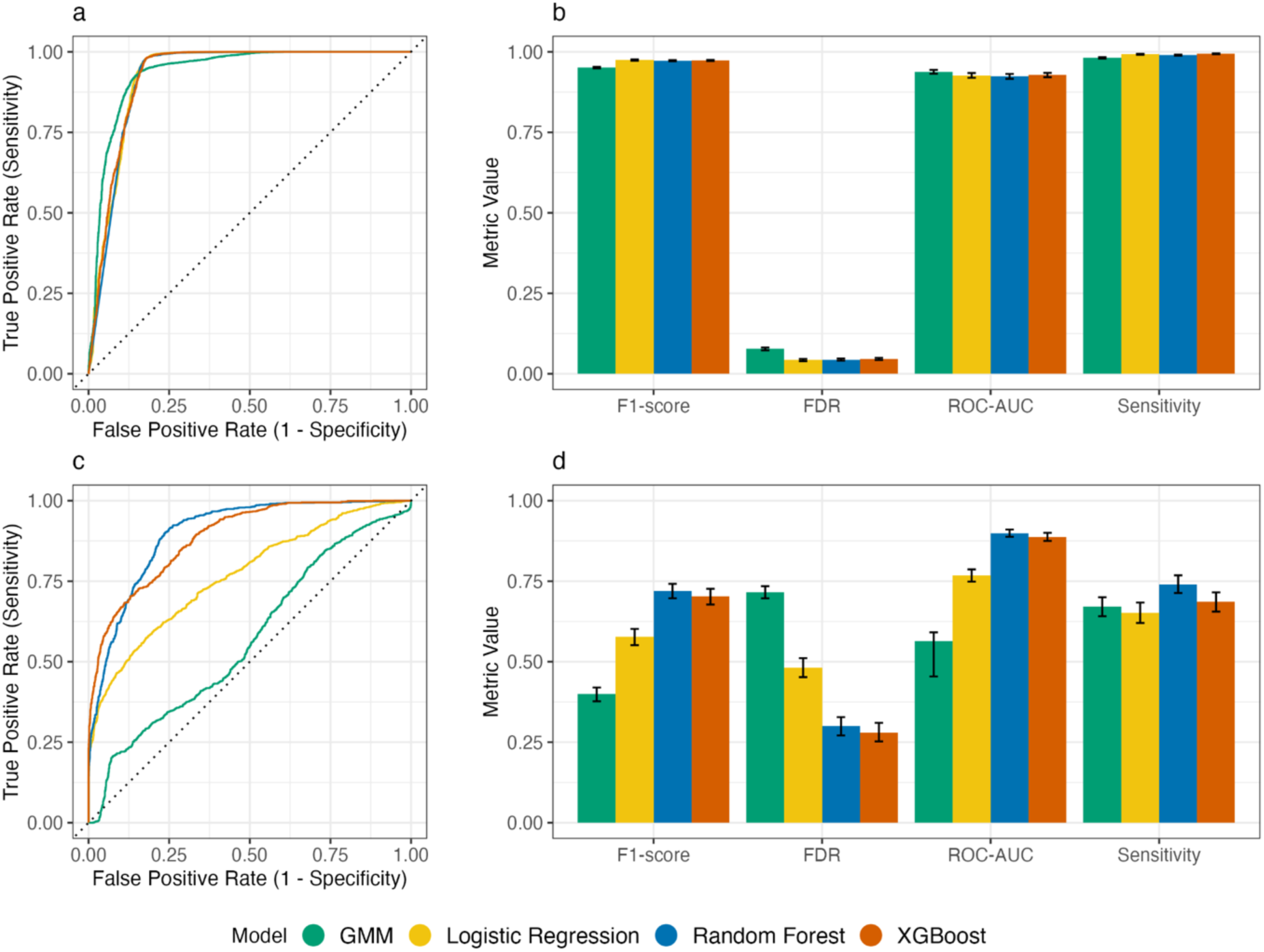
Performance of four variant filtering models in a held-out test set. (**a-b**) Performance of models trained and tested on SNVs, and (**c-d**) performance of indel models. (**a,c**) ROC curves. (**b,d**) Performance metrics. Error bars indicate the 95% confidence interval calculated using bootstrapping.

### Machine learning based filtering improved performance in the benchmark using the held-out dataset

To evaluate the final filtering module, the machine learning models (optimized for either F1 score or sensitivity) were benchmarked against GATK’s two recommended filtering methods: hard filtering thresholds and Variant Quality Score Recalibration (VQSR). We utilized unfiltered calls from GATK joint and individual genotyping workflows as a baseline. Again, the GATK joint genotyping workflow exhibited systematic failures in merging sample genotype likelihoods, requiring manual removal of 8 loci for the workflow to complete. This technical limitation rendered the joint genotyping workflow unsuitable for routine analysis of pooled data. For all variants in the test matrix, GATK individual genotyping showed higher sensitivity than joint genotyping (0.958 vs. 0.923) with comparable FDR (0.255 vs. 0.252) (see **Figure 3**). The F1-optimized ML filter achieved the most substantial FDR reduction (0.25 to 0.06) while maintaining high sensitivity (0.958 to 0.935). In comparison, hard filtering severely impacted sensitivity (dropping to 0.58), while the sensitivity-optimized ML model maintained its sensitivity (0.949) with an FDR reduction higher than VQSR (0.134 vs. 0.187). These results demonstrate that the machine learning-based filtering approach can improve performance compared to GATK’s recommended filtering methods in pooled sequencing data, achieving FDR reduction while maintaining high sensitivity.

**Figure 3.**
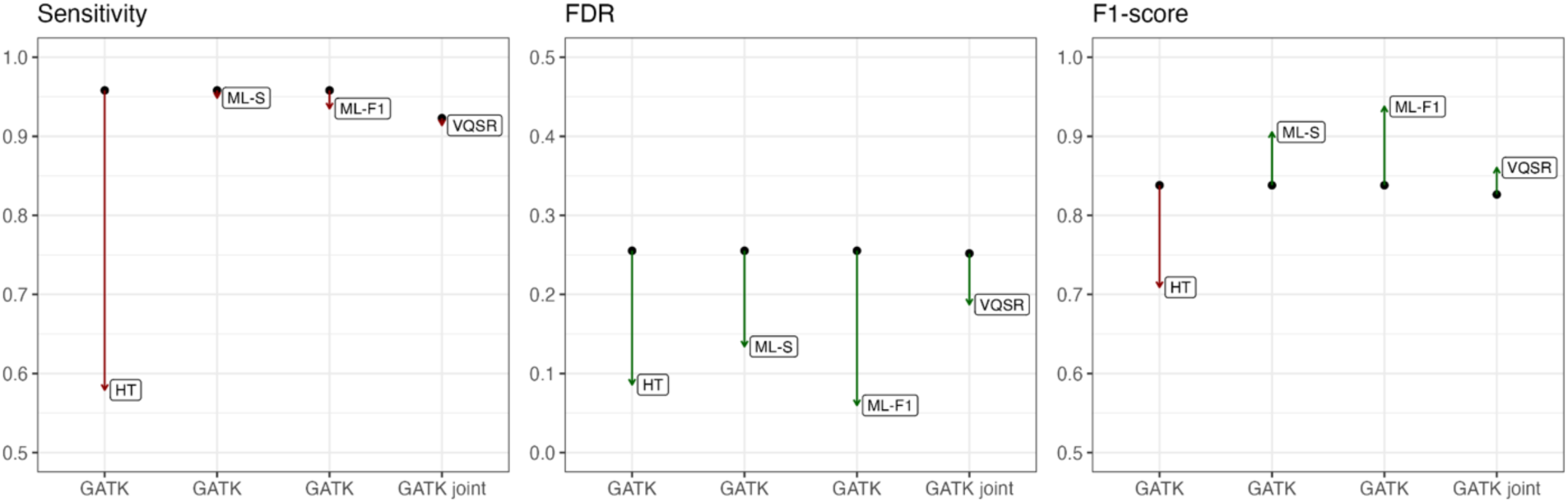
Effect of filtering approach on variant calling performance. Gain or loss of performance after filtering variant calls from GATK individual genotyping (ML-S/F1, hard filtering thresholds) or joint genotyping (VQSR) workflows.

### Filtering had a limited positive effect on pinpointable indels in comparison to SNVs

When separating pinpointable variants into SNVs and indels, differences in performance became evident (see **Figure 4**). For SNVs, the individual genotyping baseline F1-score was 0.912. The F1-optimized ML filter increased the score by 0.018 (to 0.930), and the sensitivity-optimized ML filter increased it by 0.016 (to 0.928). In contrast, VQSR only increased the joint genotyping F1-score by 0.004 (from 0.928 to 0.932). For indels, the individual genotyping baseline F1-score was 0.484 with an FDR of 0.464. Neither ML filtering approach improved the F1-score, which decreased to 0.250 (ML-F1) and 0.364 (ML-S). The joint genotyping workflow achieved a higher baseline F1-score of 0.526 with an FDR of 0.348, with VQSR having limited effect on the F1-score (0.531).

**Figure 4:**
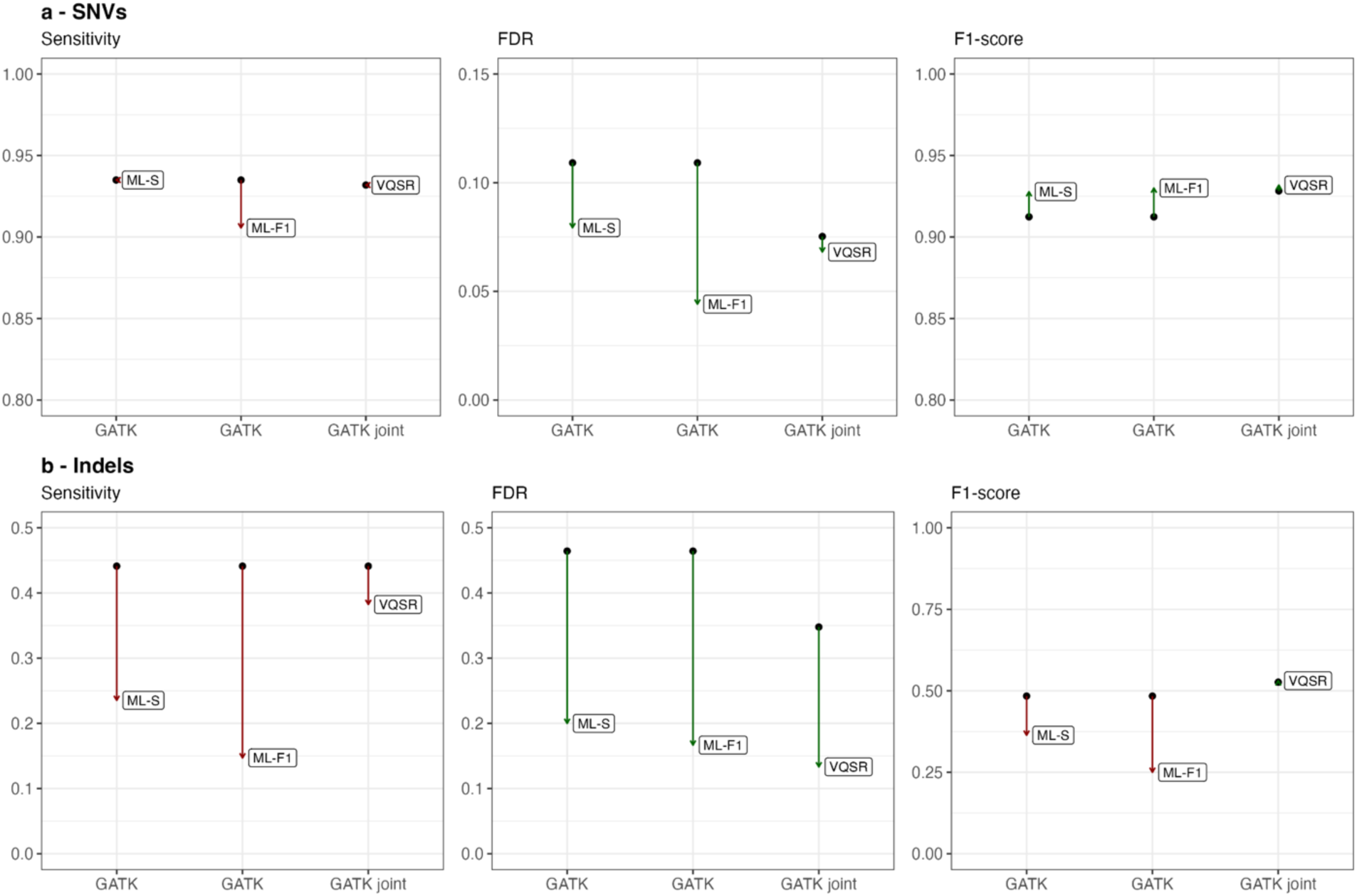
Effect of filtering approach on variant calling performance for pinpointable SNVs and indels. Gain or loss of performance after filtering pinpointable variant calls from GATK individual genotyping (ML-S/F1) or joint genotyping (VQSR) workflows. The joint genotyping workflow failed to analyse all pools. Filtering using hard thresholds was omitted due to a very large performance loss.

These results indicate that the optimal variant filter in pooled sequencing data requires different strategies for SNVs and indels. For SNVs, both ML-based approaches yield a greater increase in F1-scores compared to GATK’s recommended methods. In contrast, indel calls benefit from filtering at the pool level, while pinpointable indels only show limited improvements in the joint genotyping workflow.

## Discussion

Here we present an automated workflow for analyzing and assigning rare variants to individuals in overlapping pooled sequencing data. Using a Danish childhood cancer cohort with known genetic variation, we benchmarked multiple variant callers in the workflow and found a generally high sensitivity accompanied by high FDR. Based on GATK technical annotations, we developed a machine learning based variant filter module. A benchmark compared to hard filtering thresholds and GATK VQSR revealed higher F1 scores for the module as well as a general reduction of FDR while maintaining high sensitivities. The effect of the filter on individually pinpointed variants was less definitive, with an increasing F1 score for SNV calls but no improvement for indels.

The reported performance of variant callers in low frequency and pool-seq data is inconsistent across studies, with callers showing highly variable performances (Huang *et al*. 2015; Xiang *et al*. 2023; Maruzani *et al*. 2024). This inconsistency likely stems from differences in parameter optimization and the varying use of synthetic or real experimental data. Nevertheless, the slightly superior performance of GATK in our study is consistent with in-silico results reported by Huang et al. (2015). Furthermore, we find that CRISP did not outperform other callers in our study, contrary to previous findings(Jakaitiene, Avino and Guarracino 2017) and its use in multiple pool-seq studies (Ryu *et al*. 2018; Wei *et al*. 2020; Zhu *et al*. 2020). Although resource usage was not a primary focus of our comparison, it is noteworthy that both GATK joint genotyping workflow and Octopus failed to analyze the data successfully. In contrast, CRISP executed rapidly, which could be advantageous for larger matrix sizes.

Very few studies have investigated the performance of variant calling in overlapped pooled sequencing data, with experimental data validated against individual sequencing. One such study (Zuzarte *et al*. 2014), which employed a design similar to DoBSeq, randomly sampled 60 variants in a 12×12 matrix of individuals and verified these by individual sequencing. This approach did not account for false negative calls, making it impossible to determine the sensitivity of their method. Nevertheless, they reported FDRs of 0.1 and 0.2 for all callable and pinpointable variants, respectively. These results are converse to the FDRs of 0.2 and 0.1 observed with raw calls in our workflow. In our case, the FDR decreased, as expected, when assessing only pinpointed variants since the algorithm inherently applies additional filtering by requiring a variant to be detected in two pools. The accompanying decrease in sensitivity for pinpointable variant calls is likely attributable to their lower allelic frequency in the pools. Most variants in the call set are shared among multiple individuals in each pool, resulting in higher frequencies, making them readily detectable with a mean coverage of more than 3000X in most pools. Nonetheless, compared to the study above, our method achieved superior FDRs of 0.06 and 0.04 after applying machine learning filtering.

The binary classification task of filtering true and false variant calls in our analysis proved substantially easier for SNVs than indels. All model types performed strongly in filtering SNVs, achieving high sensitivity (0.98-0.99) and low false discovery rates (0.04- 0.08) with F1-optimized thresholds. In contrast, indel filtering sensitivities ranged from 0.65 to 0.74, and FDRs from 0.3 to 0.74. This difference is likely partly caused by the much smaller indel training dataset (2126 vs. 12761) with significantly lower confidence in its labeling. In the gold standard dataset, 32% (685/2126) of indels were called by only one of the two callers (GATK or DeepVariant), a discordance that aligns with findings from other studies (Supernat *et al*. 2018; Lin *et al*. 2022). Such a smaller and noisier dataset inherently creates worse conditions for the models to learn patterns in the technical annotations and generalize effectively.

Another reason for the differential challenge of classifying SNVs and indel errors is likely their different origins. False positive SNVs often arise due to substitutions during sequencing and incorrect alignment (Li 2014; Ma *et al*. 2019). Both cases should theoretically produce discriminatory quality metrics that can be addressed through filtering models like the one developed here. Although overlapping, the primary source is likely slightly different for indels. In a study by Li et al. (2014), low-complexity regions (LCRs) were shown to harbor 80-90% of indel errors despite comprising only 2% of the genome. They identify two main sources of errors in these regions: PCR amplification artifacts, often along homopolymer runs, and sub-optimal alignments in LCRs that are not efficiently resolved, even with local-reassembly-based variant callers. The discordance in indel calls between GATK and DeepVariant likely stems from these sub- optimal alignments, as GATK performs local reassembly while DeepVariant does not.

These errors should, however, be discernible through quality metrics and, therefore, filterable. On the other hand, PCR amplification errors are much more challenging to detect since they occur before sequencing and consequently appear indistinguishable from true mutations, though possibly at lower frequencies. Rather than addressing this specific type of error computationally, a more direct biochemical approach might be more effective, as reviewed by Salk et al. (2018).

The primary motivation for pooled sequencing is cost reduction, acknowledging that the sensitivity will never exceed individual sequencing. While the two-dimensional pooling strategy may not minimize the number of tests per positive sample as effectively as other methods, it provides important advantages. A notable alternative approach relies on hierarchical pooling, initially using large pools and then creating progressively smaller pools from positive results (Furstenau *et al*. 2020). This method has the potential to reduce the sequencing costs even further. However, hierarchical pooling requires precise knowledge of the target variants at the time of analysis, as subsequent laboratory steps depend on initial findings. The primary advantage of the two- dimensional pooling strategy is that all rare variants are assigned in a single step. Many disease-relevant variants are of unknown significance, but their interpretation is likely to evolve with ongoing research. Our method allows variant assignment data to be easily re-analyzed against updated databases. This is particularly straightforward using an automated pipeline, where modules can be easily modified and steps rerun. Similarly, updating the workflow and rerunning the entire sequence analysis remains uncomplicated as new NGS analysis tools and variant callers are developed.

Finally, using WGS data as a benchmark comes with certain limitations. Based on the results by Sun et al. (2021), 30x WGS is likely to achieve >99% sensitivity and positive predictive value (PPV) for both homozygous and heterozygous SNVs. However, the sensitivity for indels will be lower and likely below 90% for both homozygous and heterozygous indels, with a PPV below 85% for heterozygous indels. These accuracy limitations will arguably impact the validation process, as false-positive WGS calls may be incorrectly labeled as false negatives in the variant calls from DoBSeq data, and false-negative WGS calls may lead to incorrectly labeled false positives. This challenge could be addressed by increasing WGS sequencing depth, incorporating long-read sequencing technologies, or using DNA from well-characterized reference genomes as part of the matrices.

## Data Availability

The DoBSeq workflow is publicly available at https://github.com/RasmussenLab/DoBSeqWF, and the machine learning filter models, along with scripts for reproducing data analyses and figures, can be accessed at https://github.com/madscort/dwf-filter/. The underlying sequence data cannot be shared publicly due to the privacy of individuals who participated in the study and restrictions from the ethical approval. The data will be shared on reasonable request to the corresponding author, subject to appropriate ethical and legal approvals.

## Declarations

### Ethics approval and consent to participate

This research was approved by the Capital Region Committee on Health Research Ethics (H-15016782) and the Danish Data Protection Agency (RH-2016-219). Written and oral consent was obtained from parents or legal guardians for each participant’s involvement in the study. In accordance with Danish Law, adolescent participants 15 years or older were actively included in the consent process. This meant that while parents/legal guardians legally make the final decision regarding the study participation of any child under 18 years of age, adolescent participants were invited into the informed consent process, ensuring that older children’s perspectives and preferences were heard and that their will was fully considered in the decision. This study adhered to the principles of the Helsinki Declaration.

### Authors’ contributions

Conceptualization (STAGING): KS, KW, UKS; Methodology (DoBSeq): JBG, UKS, HH, CMH, SR, KS, KW, TvOH; Methodology (ML filtering): MC, SR, CMH; Software: MC, CMH; Investigation: MC; Data curation: CMH, UKS; Resources: KW, JBG, MN, CMH, AB, MABH; Supervision: SR, UKS, KW; Writing – original draft: MC; Writing – review & editing: All authors

## Acknowledgments

Anja Hirche, Solvej Margarete Aldinger Kullegaard, and colleagues for coordination and executions of patient inclusion, sample collection, and data curation.

## References

Amberger JS, Bocchini CA, Schiettecatte F et al. OMIM.org: Online Mendelian Inheritance in Man (OMIM®), an online catalog of human genes and genetic disorders. Nucleic Acids Res 2015;43:D789–98.

Andrews S. FastQC: A Quality Control Tool for High Throughput Sequence Data. www.bioinformatics.babraham.ac.uk/projects/fastqc/

Bansal V. A statistical method for the detection of variants from next-generation resequencing of DNA pools. Bioinformatics 2010;26:i318–24.

Bick D, Jones M, Taylor SL et al. Case for genome sequencing in infants and children with rare, undiagnosed or genetic diseases. J Med Genet 2019;56:783–91.

Boycott KM, Rath A, Chong JX et al. International Cooperation to Enable the Diagnosis of All Rare Genetic Diseases. Am J Hum Genet 2017;100:695–705.

Chen T, Guestrin C. XGBoost: A Scalable Tree Boosting System. Proceedings of the 22nd ACM SIGKDD International Conference on Knowledge Discovery and Data Mining. New York, NY, USA: Association for Computing Machinery, 2016, 785–94.

Chung CCY, Hong Kong Genome Project, Chu ATW et al. Rare disease emerging as a global public health priority. Front Public Health 2022;10, DOI: 10.3389/fpubh.2022.1028545.

Cooke DP, Wedge DC, Lunter G. A unified haplotype-based method for accurate and comprehensive variant calling. Nat Biotechnol 2021;39:885–92.

Danecek P, Bonfield JK, Liddle J et al. Twelve years of SAMtools and BCFtools. GigaScience 2021;10:giab008.

Das S, Biswas NK, Basu A. Mapinsights: deep exploration of quality issues and error profiles in high-throughput sequence data. Nucleic Acids Res 2023;51:e75.

DePristo MA, Banks E, Poplin R et al. A framework for variation discovery and genotyping using next-generation DNA sequencing data. Nat Genet 2011;43:491–8.

Di Tommaso P, Chatzou M, Floden EW et al. Nextflow enables reproducible computational workflows. Nat Biotechnol 2017;35:316–9.

Ferreira CR. The burden of rare diseases. Am J Med Genet A 2019;179:885–92.

Furstenau TN, Cocking JH, Hepp CM et al. Sample pooling methods for efficient pathogen screening: Practical implications. PLoS ONE 2020;15:e0236849.

Haendel M, Vasilevsky N, Unni D et al. How many rare diseases are there? Nat Rev Drug Discov 2020;19:77.

Huang HW, Mullikin JC, Hansen NF et al. Evaluation of variant detection software for pooled next-generation sequence data. BMC Bioinformatics 2015;16:235.

Jakaitiene A, Avino M, Guarracino MR. Beta-Binomial Model for the Detection of Rare Mutations in Pooled Next-Generation Sequencing Experiments. J Comput Biol 2017;24:357–67.

Koboldt DC. Best practices for variant calling in clinical sequencing. Genome Med 2020;12:91.

Li H. Toward better understanding of artifacts in variant calling from high-coverage samples. Bioinformatics 2014;30:2843–51.

Li H, Durbin R. Fast and accurate long-read alignment with Burrows–Wheeler transform. Bioinformatics 2010;26:589–95.

Lin Y-L, Chang P-C, Hsu C et al. Comparison of GATK and DeepVariant by trio sequencing. Sci Rep 2022;12:1809.

Ma X, Shao Y, Tian L et al. Analysis of error profiles in deep next-generation sequencing data. Genome Biol 2019;20:50.

Maruzani R, Brierley L, Jorgensen A et al. Benchmarking UMI-aware and standard variant callers for low frequency ctDNA variant detection. BMC Genomics 2024;25:827.

McKenna A, Hanna M, Banks E et al. The Genome Analysis Toolkit: A MapReduce framework for analyzing next-generation DNA sequencing data. Genome Res 2010;20:1297–303.

Pedregosa F, Varoquaux G, Gramfort A et al. Scikit-learn: Machine Learning in Python. J Mach Learn Res 2011;12:2825–30.

Poplin R, Chang P-C, Alexander D et al. A universal SNP and small-indel variant caller using deep neural networks. Nat Biotechnol 2018;36:983–7.

Radovic M, Ghalwash M, Filipovic N et al. Minimum redundancy maximum relevance feature selection approach for temporal gene expression data. BMC Bioinformatics 2017;18:9.

Ryu S, Han J, Norden-Krichmar TM et al. Effective discovery of rare variants by pooled target capture sequencing: A comparative analysis with individually indexed target capture sequencing. Mutat Res Mol Mech Mutagen 2018;809:24–31.

Salk JJ, Schmitt MW, Loeb LA. Enhancing the accuracy of next-generation sequencing for detecting rare and subclonal mutations. Nat Rev Genet 2018;19:269–85.

Stark Z, Scott RH. Genomic newborn screening for rare diseases. Nat Rev Genet 2023;24:755–66.

Stoltze UK, Hagen CM, van Overeem Hansen T et al. Combinatorial batching of DNA for ultralow-cost detection of pathogenic variants. Genome Med 2023;15:17.

Sun Y, Liu F, Fan C et al. Characterizing sensitivity and coverage of clinical WGS as a diagnostic test for genetic disorders. BMC Med Genomics 2021;14:102.

Supernat A, Vidarsson OV, Steen VM et al. Comparison of three variant callers for human whole genome sequencing. Sci Rep 2018;8:17851.

Van der Auwera GA, Carneiro MO, Hartl C et al. From FastQ data to high confidence variant calls: the Genome Analysis Toolkit best practices pipeline. Curr Protoc Bioinforma 2013;43:11.10.1–11.10.33.

Vasimuddin Md, Misra S, Li H et al. Efficient Architecture-Aware Acceleration of BWA- MEM for Multicore Systems. 2019 IEEE International Parallel and Distributed Processing Symposium (IPDPS). 2019, 314–24.

Wei T, Simko V. R package “corrplot”: Visualization of a Correlation Matrix. 2024.

Wei YB, McCarthy M, Ren H et al. A functional variant in the serotonin receptor 7 gene (HTR7), rs7905446, is associated with good response to SSRIs in bipolar and unipolar depression. Mol Psychiatry 2020;25:1312–22.

Wilm A, Aw PPK, Bertrand D et al. LoFreq: a sequence-quality aware, ultra-sensitive variant caller for uncovering cell-population heterogeneity from high-throughput sequencing datasets. Nucleic Acids Res 2012;40:11189–201.

Wright CF, FitzPatrick DR, Firth HV. Paediatric genomics: diagnosing rare disease in children. Nat Rev Genet 2018;19:253–68.

Xiang X, Lu B, Song D et al. Evaluating the performance of low-frequency variant calling tools for the detection of variants from short-read deep sequencing data. Sci Rep 2023;13:20444.

Zhu S, He M, Liu Z et al. Shared genetic susceptibilities for irritable bowel syndrome and depressive disorder in Chinese patients uncovered by pooled whole-exome sequencing. J Adv Res 2020;23:113–21.

Zuzarte PC, Denroche RE, Fehringer G et al. A Two-Dimensional Pooling Strategy for Rare Variant Detection on Next-Generation Sequencing Platforms. PLOS ONE 2014;9:e93455.

