## Supplementary material for "DoBSeqWF: A framework for sensitive detection of individual genetic variation in pooled sequencing data"

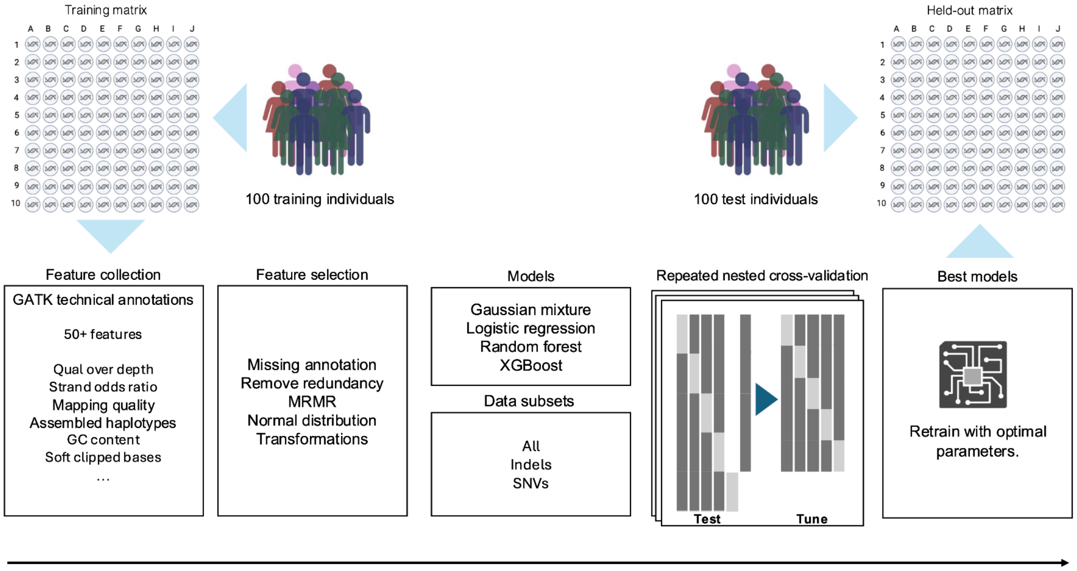


**Supplementary Figure 1.** Filter model training overview. The models are trained and tested on separate 10x10 DoBSeq matrices each including data from 100 individuals. All feature selection, hyperparameter tuning and final model selection is done using the variant data from the training set. The final prediction results and benchmark is performed with saved model weights on the full held-out test set.

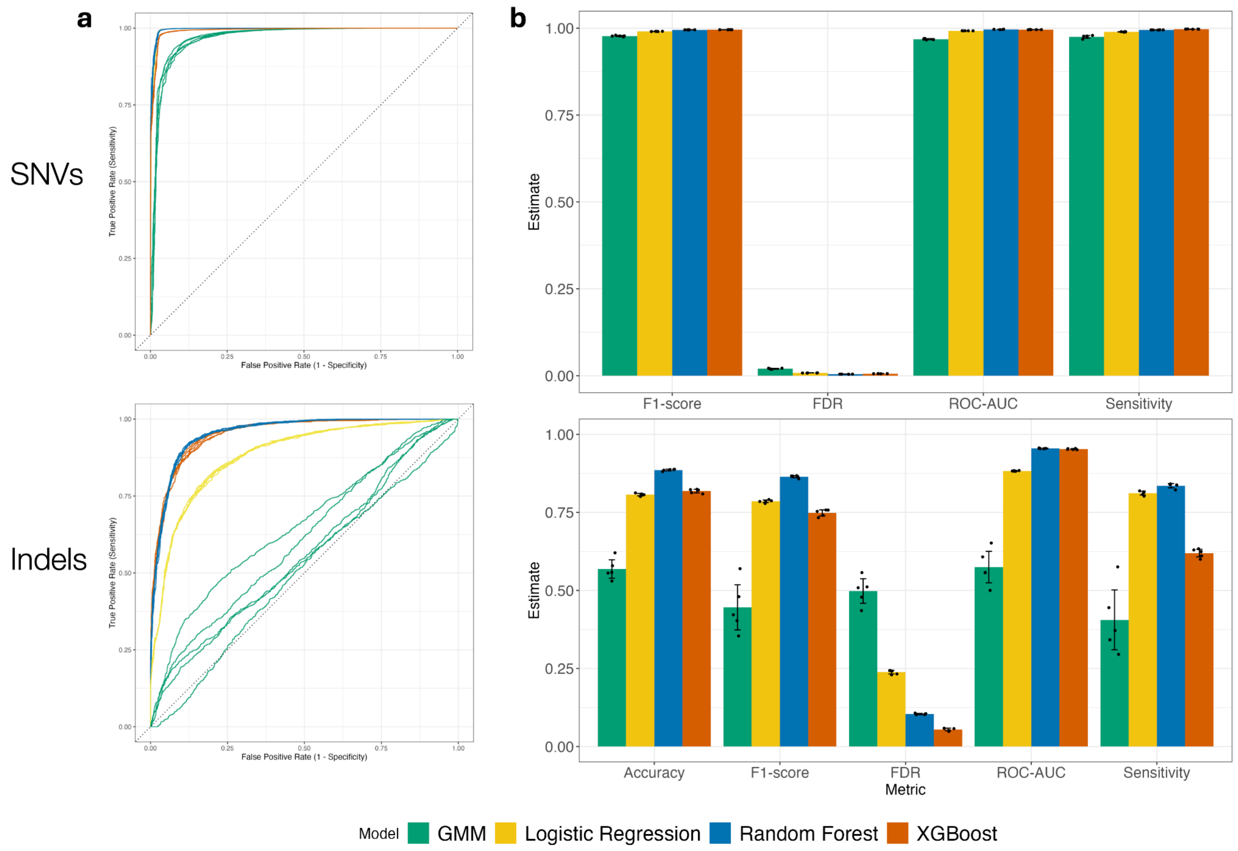


**Supplementary Figure 2.** Performance of four variant filtering models during training using repeated nested cross validation. (**a-b**) Performance of models trained and tested on SNVs, and (**c-d**) performance of indel models. (**a,c**) ROC curves with one line per repetition. (**b,d**) Performance metrics. Height indicates the mean over all repetitions, points indicate result per repetition and error bars indicate the 95% confidence interval of the mean.

| Type | Feature ID | Description |
|---|---|---|
| Site-specific | GQ | Genotype quality |
| Site-specific | DP | Approximate read depth; some reads may have been filtered |
| Site-specific | ASSEMBLED_HAPS | Haplotypes detected by the assembly region before haplotype filtering is applied |
| Site-specific | HAPCOMP | Edit distances of each alt allele's most common supporting haplotype from closest germline haplotype, excluding differences at the site in question |
| Site-specific | HAPDOM | For each alt allele, fraction of read support that best fits the most-supported haplotype containing the allele |
| Site-specific | HEC | Counts of support for haplotype groups excluding difference at the site in question |
| Site-specific | MLEAC | Maximum likelihood expectation (MLE) for the allele counts (not necessarily the same as the AC), for each ALT allele, in the same order as listed |
| Site-specific | MLEAF | Maximum likelihood expectation (MLE) for the allele frequency (not necessarily the same as the AF), for each ALT allele, in the same order as listed |
| Site-specific | X_GCC | Flow: percentage of G or C in the window around hmer |
| Allele-specific | AF | Allele Frequency, for each ALT allele, in the same order as listed |
| Allele-specific | AD | Allelic depths for the ref and alt alleles in the order liste |
| Allele-specific | BaseQRankSum | Allele specific Z-score from Wilcoxon rank sum test of each Alt Vs. Ref base qualities] |
| Allele-specific | FS | Phred-scaled p-value using Fisher's exact test to detect strand bias |
| Allele-specific | MQ | RMS Mapping Quality |
| Allele-specific | MQRankSum | Z-score From Wilcoxon rank sum test of Alt vs. Ref read mapping qualities |
| Allele-specific | QD | Variant Confidence/Quality by Depth |
| Allele-specific | ReadPosRankSum | Z-score from Wilcoxon rank sum test of Alt vs. Ref read position bias |
| Allele-specific | SOR | Symmetric Odds Ratio of 2x2 contingency table to detect strand bias |
| Allele-specific | ClippingRankSum | Z-score From Wilcoxon rank sum test of Alt vs. Ref number of hard clipped bases |
| Allele-specific | LikelihoodRankSum | Z-score from Wilcoxon rank sum test of Alt Vs. Ref haplotype likelihoods |

**Supplementary Table 1.** Description of technical annotations used for the final filtering models as given by GATK. Annotations are either site or variant allele specific.

| Model Type | Parameter | SNV range | Indel range |
|---|---|---|---|
| GMM | n_components | 2-8 | 2-8 |
| GMM | covariance_type | full, tied | full, tied |
| Logistic Regression | C | 0.00001-100 | 0.00001-500 |
| Logistic Regression | penalty | l1, l2, elasticnet | l1, l2, elasticnet |
| Logistic Regression | class_weight | None, balanced | None, balanced |
| Random Forest | n_estimators | 50-500 | 5-250 |
| Random Forest | max_depth | 5-30 | 5-30 |
| Random Forest | min_samples_leaf | 2-10 | 2-10 |
| Random Forest | class_weight | None, balanced | None, balanced |
| XGBoost | n_estimators | 10-300 | 10-300 |
| XGBoost | max_depth | 3-10 | 3-10 |
| XGBoost | learning_rate | 0.01-0.9 | 0.01-0.9 |
| XGBoost | colsample_bytree | 0.6-1.0 | 0.6-1.0 |
| XGBoost | class_weight | None, balanced | None, balanced |

**Supplementary Table 2.** Hyperparameter search ranges for the model types used in the study.
